# Exploring Temporal Dynamics in No-Reflow Assessment

**DOI:** 10.64898/2025.12.08.25341865

**Authors:** Carlos Andres Olivares Reboredo, Shreeram Athreya, Ameera Ismail, Kambiz Nael, William Speier, Corey Arnold

## Abstract

**Background:** For acute ischemic stroke (AIS) patients, successful endovascular thrombectomy (EVT) may not recover functional independence. One potential mechanism driving this disparity is no-reflow, a lack of perfusion despite recanalization of the large vessel occlusion (LVO). None of the published methods for assessing no-reflow have gained acceptance, and debate persists about isolating no-reflow from upstream hemodynamic anomalies as well as the temporal dynamics no-reflow assessment. This work conducts an exploratory analysis of the effect of time since EVT on the performance of different methods for assessing no-reflow.

**Methods:** We identified a cohort of UCLA Health patients with successful LVO recanalization (≥ mTICI 2b) of a thrombus located in M1 or ICA. Patients were stratified into ‘early’ and ‘follow-up’ cohorts of < *24* and < *48* hours since EVT. A set of published methods using perfusion imaging for no-reflow detection were adapted to run semi-automatically for both cohorts.

**Results:** We compiled 83 imaging studies from 64 patients. No-reflow rates varied from *4*.*2*% to *51*.*4*% by method, time since EVT, and mTICI. Follow-up no-reflow rates more closely match the original literature across all methods. mTICI 2b patients likely inflated no-reflow, as rates dropped approximately *50*% for mTICI 2c/3 cases. Early cohort inter-method agreement shows kappa scores as low as −*0*.*017*, but improved up to *0*.*72* in the follow-up cohort.

**Conclusion:** No-reflow exhibits substantial dependence on time across all no-reflow detection methods studied. Inter-method agreement is good for methods using the same perfusion parameters, but each method is subject to confounding influences, motivating an aggregate method for robust evaluation. The temporal dynamics of no-reflow detection raises questions about imaging timing post-EVT for detection within the intervention window.

## Introduction

Stroke kills roughly 190,000 people in the United States every year [1]. Of the roughly 795,000 annual stroke cases in the United States, 87% are ischemic strokes, making timely and effective interventions imperative [2]. Physicians rely on endovascular thrombectomy (EVT) as a first line therapy for the treatment of large vessel occlusion (LVO) acute ischemic stroke (AIS) [3], [4]. EVT success is typically measured via the thrombolysis in cerebral infarction (TICI) scale, which ranges from 0 (no recanalization) to 3 (complete antegrade recanalization of ischemic territory) [5]. Subsequent scales such as the extended (eTICI) and modified (mTICI) were developed to refine the 2a and 2b scores of the original method. Intervention with EVT is generally considered a success if patency is restored to the occluded vessel at the level of mTICI 2b, eTICI 2b50, or better [6].

Researchers and clinicians alike observe that success in EVT does not necessarily translate to success in recovering functional independence for patients [7], [8], [9]. In particular, patients whose occlusions are recanalized via EVT have a higher chance of achieving a good functional outcome (a modified Rankin score (mRS) ≤ *2* at 90 days post-intervention), but a significant number of patients do not meet this mRS threshold even if their procedure was scored as mTICI 2c or 3 [10], [11]. One mechanism driving these anomalies is a lack of reperfusion of the cerebral parenchyma despite recanalization of the occluded vessel, a phenomenon termed no-reflow [12]. No-reflow represents a new target in AIS treatment, as developing preventative treatments or interventions for this post-EVT complication would improve patient outcomes [13].

Accurately characterizing microcirculatory pathologies (no-reflow) in the brain is wrought with unique challenges for both intra- and peri-interventional imaging. Digital subtraction angiography (DSA) images taken during EVT require longer x-ray pulse durations and lower frame rates than other angiographic procedures (3 fps compared to 15-30 fps for percutaneous coronary intervention), removing the possibility of using blush frames to estimate reperfusion status as in myocardial blush grade for cardiac ischemia [14], [15]. Cerebral tissue also displays a high degree of heterogeneity and has a complex vascular structure, leading to individual pixels or voxels capturing the hemodynamics of both no-reflow (ischemic) and perfused tissue. Collateral flow further adds noise to signal [16]. The combined effect of these challenges limits healthcare practitioners’ ability to directly observe cerebral no-reflow with computed tomography (CT) or magnetic resonance (MR).

Perfusion imaging provides clinicians with reliable, if indirect, methods of estimating tissue reperfusion in the brain without microscopic imaging. During perfusion weighted imaging, a sequence of 3-dimensional scans is acquired over time, capturing the dynamics of an intravascular tracer. From these 4-dimensional sequences, processing methods such as deconvolution estimate perfusion parameters such as the relative cerebral blood volume or flow (rCBV or rCBF), mean transit time (MTT), time to the maximum of the residue function (Tmax), and more [17]. Perfusion imaging is possible with both MR and CT, making it a flexible option for many institutions. Moreover, studies like the DAWN [18], DEFUSE 3 [19], and EXTEND [20] trials have shown that perfusion imaging can be leveraged to make treatment decisions, and have contributed thresholds on different parameters to classify tissue as hypoperfused. In fact, there is a broad range of work describing post-intervention perfusion abnormalities using different thresholds on parameters to identify no-reflow with perfusion imaging [21], [22], [23], [24].

No generally accepted method exists for assessing no-reflow with clinically available perfusion imaging. Current approaches characterize the phenomenon with different parameters, post-processing techniques, and patient selection criteria, incurring trade-offs between simplicity and accuracy. A Tmax > 6 s threshold reliably identifies critically hypoperfused tissue in major clinical trials [25]. Tissue optimal reperfusion (TOR), defined as a reduction in the brain volume with Tmax > 6 s by 90% or more between pre- and post-EVT perfusion, serves as a no-reflow detection method, correlating strongly with the final volume of hypoperfused tissue [23], [26]. Other studies report that rCBV and rCBF reductions of ≤ 30% relative to the contralateral region identify irreversibly damaged tissue, with a decrease to ≤15% compared to unaffected tissue as a lower bound for evaluating hypoperfusion consistent with no-reflow [17], [24].

Debate persists on recanalization thresholds sufficient for isolating microvascular dysfunction, typically defined by modified TICI (mTICI) or expanded TICI (eTICI) grades [27], [28]. Although mTICI ≥ 2b is often deemed adequate reperfusion, residual macrovascular flow deficits may confound microvascular assessment, prompting some to restrict analysis to mTICI 2C–3 cases. Inconsistencies in mTICI grading across raters and institutions further complicate inclusion, often requiring labor-intensive consensus re-scoring [29].

Another noted gap in the literature is a clear understanding of how the timing of follow-up imaging may influence the performance of each proposed no-reflow detection method [22]. Existing research makes use of either clinical trial data with rigid imaging windows or single-institution retrospective cohorts with highly variable protocols. This dichotomy of data sources results in a disjointed research landscape where the imaging window for detecting no-reflow ranges from within 30 minutes to almost 72 hours post-EVT [22], [23], [24], [30] and no one study is well equipped to study the influence of time since EVT on no-reflow detection.

The object of our work is to assess no-reflow according to multiple published methods and analyze how they depend on time since EVT using ‘early’ and ‘follow-up’ cohorts of patients treated in a clinical setting, as well as patients with multiple post-EVT imaging instances. In pursuit of our analysis, we implemented semi-automated computational pipelines and compared no-reflow rates as computed by our method with the rates reported in the original literature. We also discuss factors underlying the degree of agreement between the methods and possible confounding influences each method may be subject to.

## Methods

### Patient Cohort

For our analysis we identified a retrospective cohort from patients who underwent treatment at Ronald Reagan UCLA Medical Center between the years 2011-2024. We included patients according to (1) LVO AIS treated with EVT (2) Successful recanalization (mTICI ≥ 2b) (3) Primary thrombus located in M1 or ICA territory (4) Baseline and follow-up imaging included MR perfusion imaging with contrast (5) Follow-up imaging occurs within 48 hours post-EVT. The study data was collected and de-identified with approval from the institutional review board IRB-18-0329. No informed consent was required.

### Early, Follow-up, and Intersect Imaging cohorts

To investigate how time since EVT affects no-reflow frequency for each method, patients were stratified into an ‘early scan’ and a ‘follow-up scan’ cohort. A study was included in the ‘early cohort’ if it occurred within 12 hours of EVT, or if there was another post-EVT study at or after 24 hours and within 48 hours. A 19 patient subset of patients underwent more than one post-EVT MRI study with perfusion imaging, and are present in both the early and follow-up scan cohort. This subset is termed the ‘intersect’ cohort. The total number of scans used in our analysis is 83. The distribution of time since EVT is demonstrated in Figure 1 and an overview of the clinical variables and demographics in Table 1.

**Figure 1:**
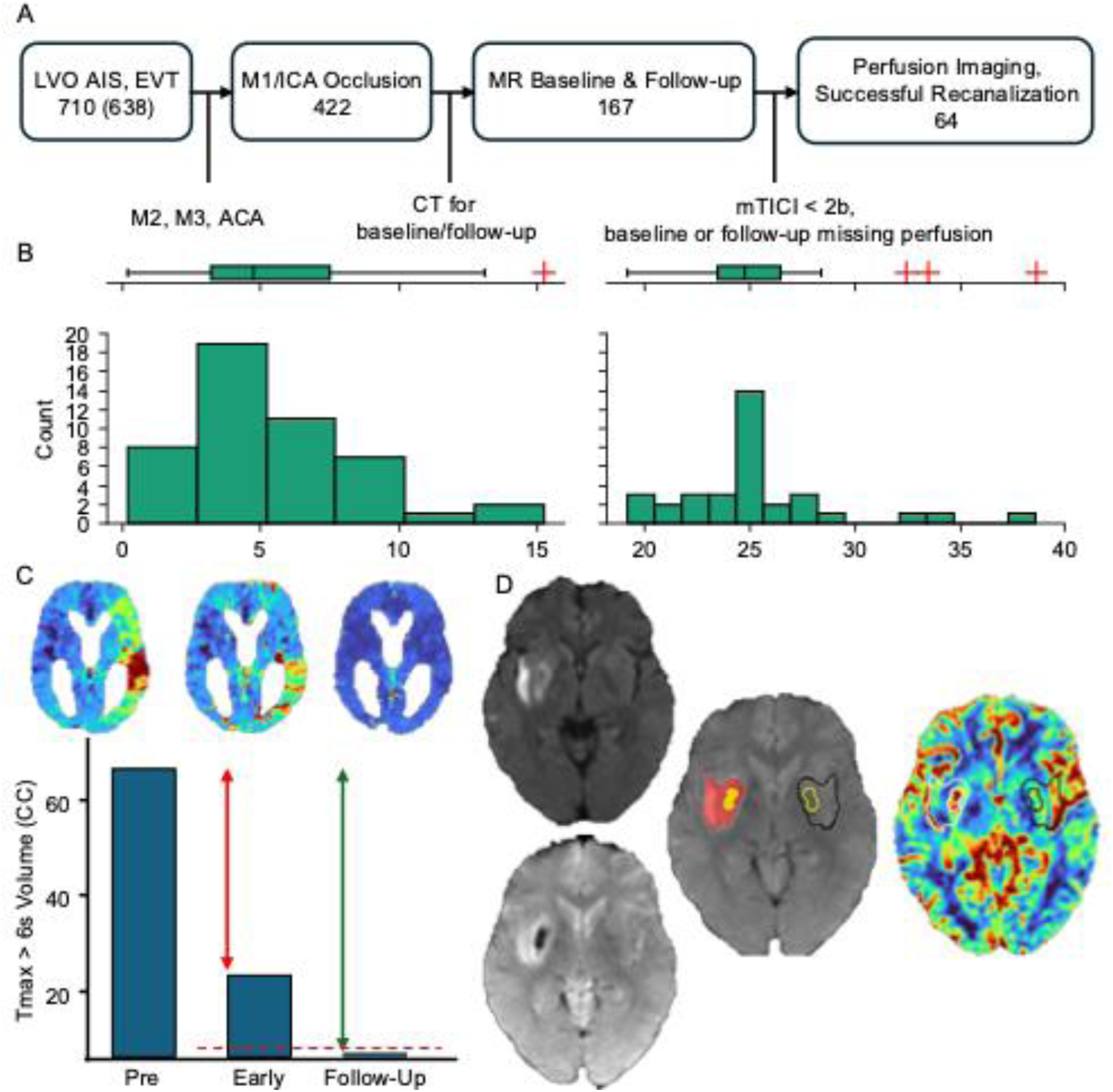
(A) Visual representation of inclusion and exclusion criteria for our analysis. UCLA is an MR-first institution. (B) Histograms of time from EVT to follow-up imaging per cohort. The most frequent time for follow-up imaging is roughly 24 hours post-EVT. An imaging study is included in the ‘early cohort’ if it occurred within 12 hours of EVT, or if there was another post-EVT study at or after 24 hours and within 48 hours. Our criteria yielded 64 unique patients. (C) Illustration of Tmax methods of Bai and Rubiera. Both methods make use of Tmax > 6 s volume in similar ways. Rubiera is dichotomized by a uniform 3.5cc threshold on post-intervention imaging. Bai compare % change in Tmax > 6 s volume, and if there is a reduction in volume less than 90%, the patient is said to have no-reflow. (D) Illustration of the rCBV/F method of Ng. First, an ROI is delineated with DWI hyperintensity segmentation. Areas of hemorrhagic transformation are subtracted from the ROI. If the median value of the ROI corresponds to a 15% or greater reduction relative to the median value of its contralateral homologue, then the patient is said to have no-reflow. Removal of HT regions represents the only step of our pipeline that is done manually.

**Table 1:**
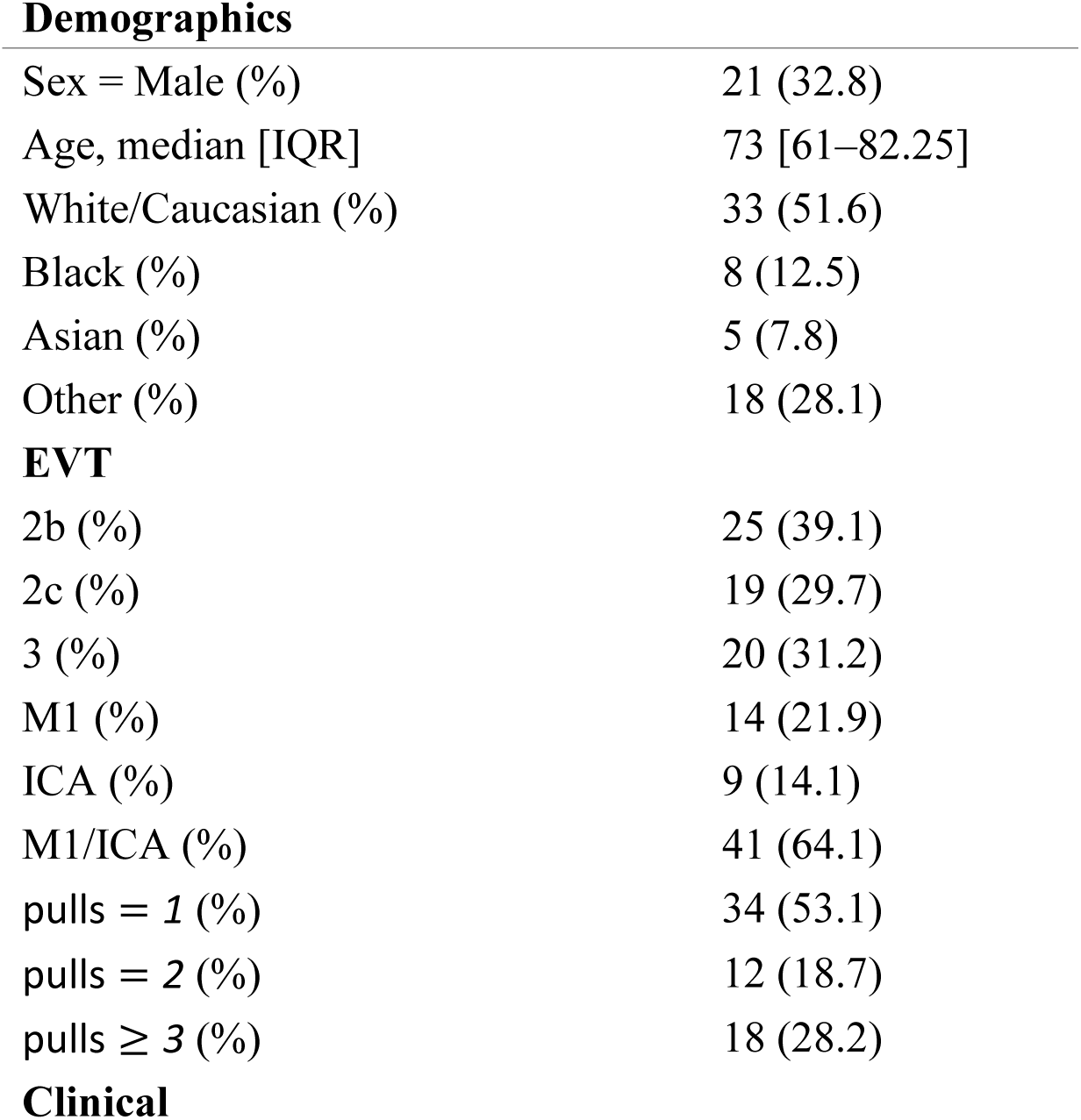
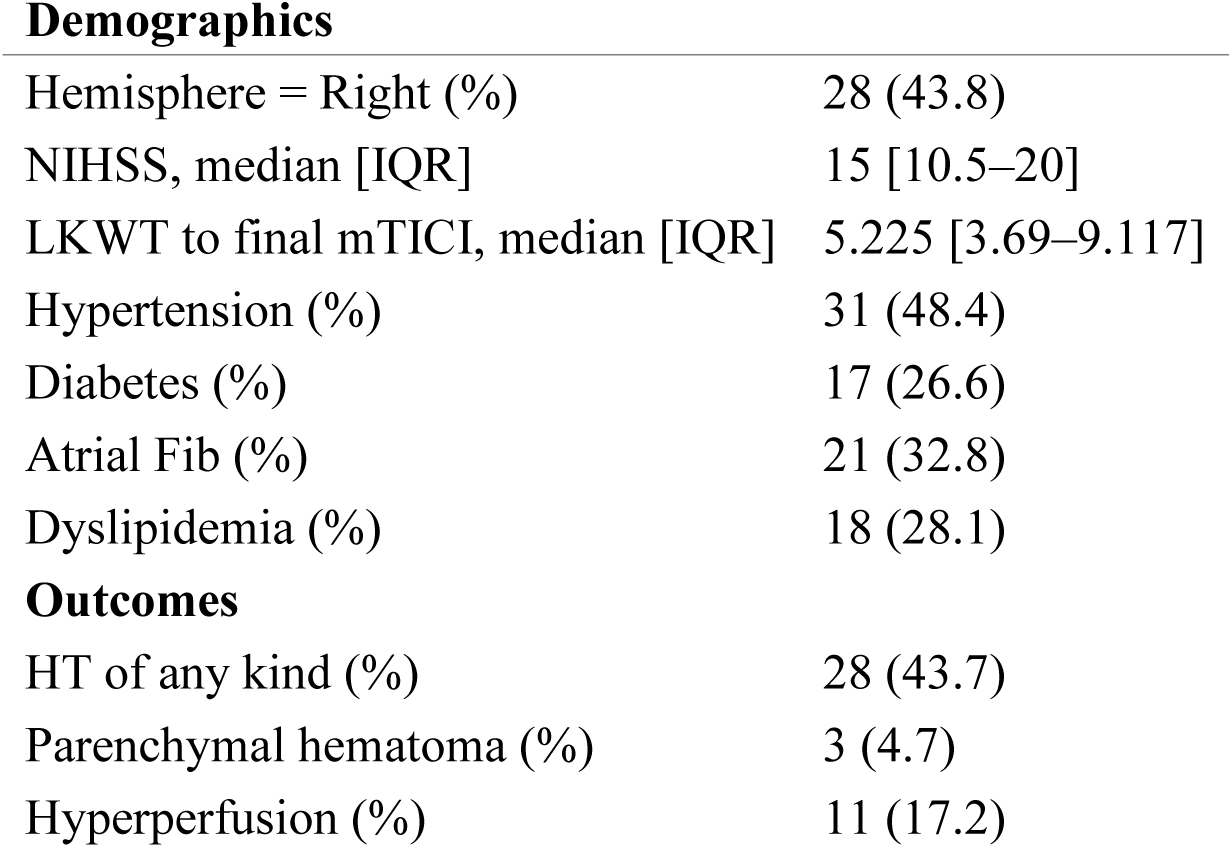
Baseline Clinical and Procedural Characteristics of the Cohort. Values are presented as median [IQR] or (n) (%). Hemorrhagic transformation (HT) confounds downstream image analysis, leading to artificially low rCBV and other artificially extreme values on other parameters, requiring masking before applying the method of Ng etal. Hyperperfusion, also referred to as luxury perfusion, also confounds downstream analysis and is the only imaging artifact that no published method accounts for.

**Demographics**
|  |  |
| --- | --- |
| Sex = Male (%) | 21 (32.8) |
| Age, median [IQR] | 73 [61–82.25] |
| White/Caucasian (%) | 33 (51.6) |
| Black (%) | 8 (12.5) |
| Asian (%) | 5 (7.8) |
| Other (%) | 18 (28.1) |
| **EVT** |  |
| 2b (%) | 25 (39.1) |
| 2c (%) | 19 (29.7) |
| 3 (%) | 20 (31.2) |
| M1 (%) | 14 (21.9) |
| ICA (%) | 9 (14.1) |
| M1/ICA (%) | 41 (64.1) |
| pulls = 1 (%) | 34 (53.1) |
| pulls = 2 (%) | 12 (18.7) |
| pulls $\geq 3$ (%) | 18 (28.2) |
| Demographics |  |
| Hemisphere = Right (%) | 28 (43.8) |
| NIHSS, median [IQR] | 15 [10.5–20] |
| LKWT to final mTICI, median [IQR] | 5.225 [3.69–9.117] |
| Hypertension (%) | 31 (48.4) |
| Diabetes (%) | 17 (26.6) |
| Atrial Fib (%) | 21 (32.8) |
| Dyslipidemia (%) | 18 (28.1) |
| Outcomes |  |
| HT of any kind (%) | 28 (43.7) |
| Parenchymal hematoma (%) | 3 (4.7) |
| Hyperperfusion (%) | 11 (17.2) |

### Imaging Protocol

Images were collected using a Siemens 1.5 T Avanto and a Siemens 3.0 T Trio. The AIS workup included diffusion weighted imaging (DWI), fluid-attenuated inversion recovery (FLAIR), dynamic susceptibility contrast MR perfusion (DSC-MRP) and gradient echo images. Perfusion parameters were generated using RAPID or OLEA softwares, including relative blood volume (rCBV), relative blood flow (rCBF), and time to peak of the residual function (Tmax).

### No-Reflow Assessment Methods

Our search was restricted to published research that proposed no-reflow detection methods with perfusion imaging. Studies were selected if they assessed macrovascular recanalization status with DSA and microvascular reperfusion status with perfusion parameters. Studies exclusively using CT were not excluded, as there is a body of evidence arguing for the interchangeable use of MR and CT perfusion imaging in AIS [31], [32]. Our study selection process is similar to that of Mutimer [22], differing mainly by omission of Schiphorst ‘s [33] method, as it was developed on arterial spin labeling (ASL) maps, which are not as frequently acquired at our institution and are difficult to evaluate in a standardized manner against methods not derived from ASL imaging.

### Extracting ROIs

To replicate Ng ‘s image processing [24] with automated methods, we developed a UNet-based model to segment the ischemic lesion given DWI and apparent diffusion coefficient (ADC) imaging. The model is first pre-trained on the public BRATS21 dataset designed for the segmentation of glioblastomas [34]. We prepared a version of the data with T2 and contrast enhanced T1 imaging. The model was then finetuned specifically for ischemic lesion segmentation with the ISLES 2022 dataset, [35] which provides DWI, ADC, and FLAIR images for segmenting such lesions from both acute and sub-acute stroke. For finetuning, we followed the method of the ‘SEALS’ team, which achieved the top rank in the ISLES 2022 Challenge [36].

### Registration, Image Processing

For each patient, a copy of the MNI152 linear atlas with spatial resolution 2mm was resampled to match the voxel dimensions of the follow-up study. This atlas will serve as the target for all downstream registrations, ensuring all DWI and perfusion sequences are co-registered to the same space. The DWI and DSC-MRP sequences were skull stripped with the HD-BET algorithm to improve registration quality [37]. Registrations were conducted with the Oxford Centre for Functional Magnetic Resonance Imaging of the Brain’s (FMRIB) Software Library’s (FSL) Linear Registration Tool (FLIRT) [38]. To register the parameter maps to the atlas, the first timepoint of the 4D DSC-MRP was extracted and registered to the custom atlas. Then, the transformation generated from the registration was applied to the rCBV, rCBF, and Tmax parameter maps. Similarly, the DWI images were registered to the custom atlas and the transformations generated were applied to the ADC sequence. This results in images that have been minimally resized and also coregistered, allowing filtering by hemisphere and comparing an ROI to its contralateral area during downstream analysis. The MNI152 linear atlas includes a ventricle mask, enabling automated removal of ventricles from the parameter maps. All registrations were affine.

### Determining No-Reflow Status

#### rCBV/F Methods

We use Ng ‘s method of classifying a > *15*% asymmetry in rCBV or rCBF in the infarct region of interest (ROI) as no-reflow. The ROI is taken from the segmented DWI hyperintensity on follow-up imaging. The median rCBV and rCBF values of this ROI are sampled and compared to the contralateral homologue, which is determined by flipping the ROI along the longitudinal fissure. If for either rCBV or rCBF the median of the affected side’s ROI is 15% or more reduced in comparison to the contralateral ROI, the patient is said to have no-reflow. Areas of confluent hemorrhagic transformation or hematomas were subtracted from the ROI before comparing medians.

#### Tmax Methods

Bai describes tissue reperfusion status with TOR and has been used in other studies as a no-reflow detection method [23], [26], [39]. Our only deviation from Bai’s original method is automatic removal of artifacts in the Tmax map compared to manual removal. This is done with filters that remove small regions of Tmax > 6 s that are disconnected from the main Tmax > 6 s lesion. Similarly, Rubiera selected patients for adjuvant treatment based on the presence of a hypoperfused (Tmax > 6 s) region with volume larger than 3.5cc on post-EVT imaging [30] and has been used for assessing no-reflow in multiple studies [22], [40]. Similarly to the method of Bai, we deviate from Rubiera’s method only with automated removal of Tmax artifacts.

## Results

For the early scan cohort, no-reflow rates varied from 4.2% to 43.8% depending on method and 2b inclusion/exclusion. For the follow-up scan cohort, rates varied from 14.3% to 51.4%. There is higher concordance in no-reflow rates between the follow-up cohort and the original literature, as shown in Table 2. There is also a uniform increase in the number of no-reflow cases in the follow-up cohort when compared to early cohort.

**Table 2:** No-reflow frequencies under four time-point definitions. ‘Mixed-Early/Late’ refers to the 64 patient cohort with the early or later follow-up studies used for the 19 intersect patients, respectively. There is a uniform increase in no-reflow occurrence across all methods for later post-intervention imaging.

|  | rate | Mixed-Early |  | Mixed-Late |  | Early scans<br>(N=48) |  | Follow-up<br>scans (N=35) |  | Mutimer |
| --- | --- | --- | --- | --- | --- | --- | --- | --- | --- | --- |
|  |  | 2b+ | 2c+ | 2b+ | 2c+ | 2b+ | 2c+ | 2b+ | 2c+ |  |
| 3-4 (lr)5-6<br>(lr)7-8 (lr)9-10 Study |  |  |  |  |  |  |  |  |  |  |
| Bai | 0.301 | 0.328 | 0.333 | 0.344 | 0.359 | 0.229 | 0.146 | 0.429 | 0.229 | 0.099 |
| Rubiera | 0.480 | 0.500 | 0.435 | 0.516 | 0.436 | 0.438 | 0.229 | 0.514 | 0.229 | 0.107 |
| Ng | 0.253 | 0.170 | 0.153 | 0.219 | 0.179 | 0.104 | 0.042 | 0.314 | 0.143 | 0.221 |

### Agreement between Methods

Agreement between methods exhibits a high degree of variability, ranging from -0.017 to 0.715. As seen in Figure 2, inter-method agreement is generally lower in the early scan cohort with average kappa score of 0.220 compared to 0.499 in the follow-up cohort. The methods of Rubiera and Bai have the highest agreement, with 0.553 in the early scan cohort and 0.715 in the follow-up cohort. The method of Ng generally exhibits poorer agreement with Tmax-based methods, with Cohen’s kappa as low as -0.017 when evaluating agreement with Rubiera’s method in the early scan cohort. However, in the follow-up scan cohort, agreement between Ng and the Tmax methods increases to fair agreement (0.265) with Rubiera and moderate agreement (0.517) with Bai.

**Figure 2:**
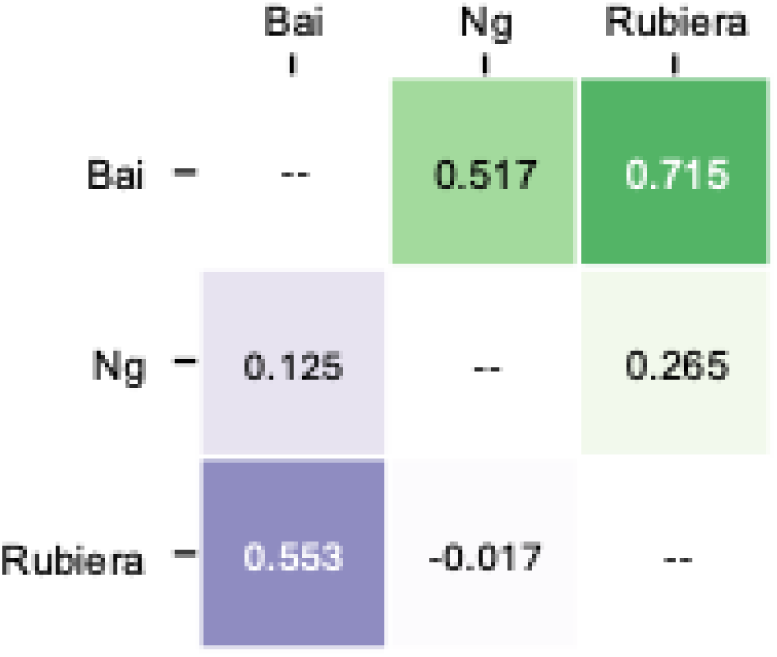
Cohen’s kappa agreement values by methods. The purple and green colormaps represent the early cohort and the follow-up cohort’s agreement values, respectively. Inter-method agreement increases uniformly in the follow-up cohort.

### Effect of 2b Inclusion/Exclusion

Across all cohorts and methods, mTICI 2b was the most-represented class of no-reflow cases, as can be seen in Figure 3. When the cohort was limited to 2c/3 cases (‘2c+’), the angiographic-time definitions (‘Mixed-Early’ and ‘Mixed-Late’ columns) showed only modest changes: the mean no-reflow frequency across studies fell from 0.333 to 0.307 for the original times (≈ *8*% relative reduction) and from 0.360 to 0.325 for the composite times (≈ *10* % reduction). By contrast, in the early perfusion scans acquired after EVT (N = 48) the mean frequency dropped by nearly half, from 0.257 to 0.139 (46 % relative reduction), and in the delayed follow-up scans (N = 35) it fell from 0.419 to 0.200 (52 % reduction). The largest single study-level effect was observed in Ng et al., where exclusion of 2b cases reduced early-scan no-reflow from 10.4 % to 4.2 %—a 60 % relative decrease. Thus, retaining mTICI 2b patients inflates no-reflow estimates, particularly in perfusion-based assessments. Restricting analyses to 2c/3 recanalization roughly halves the apparent incidence and may provide a cleaner view of microvascular failure.

**Figure 3:**
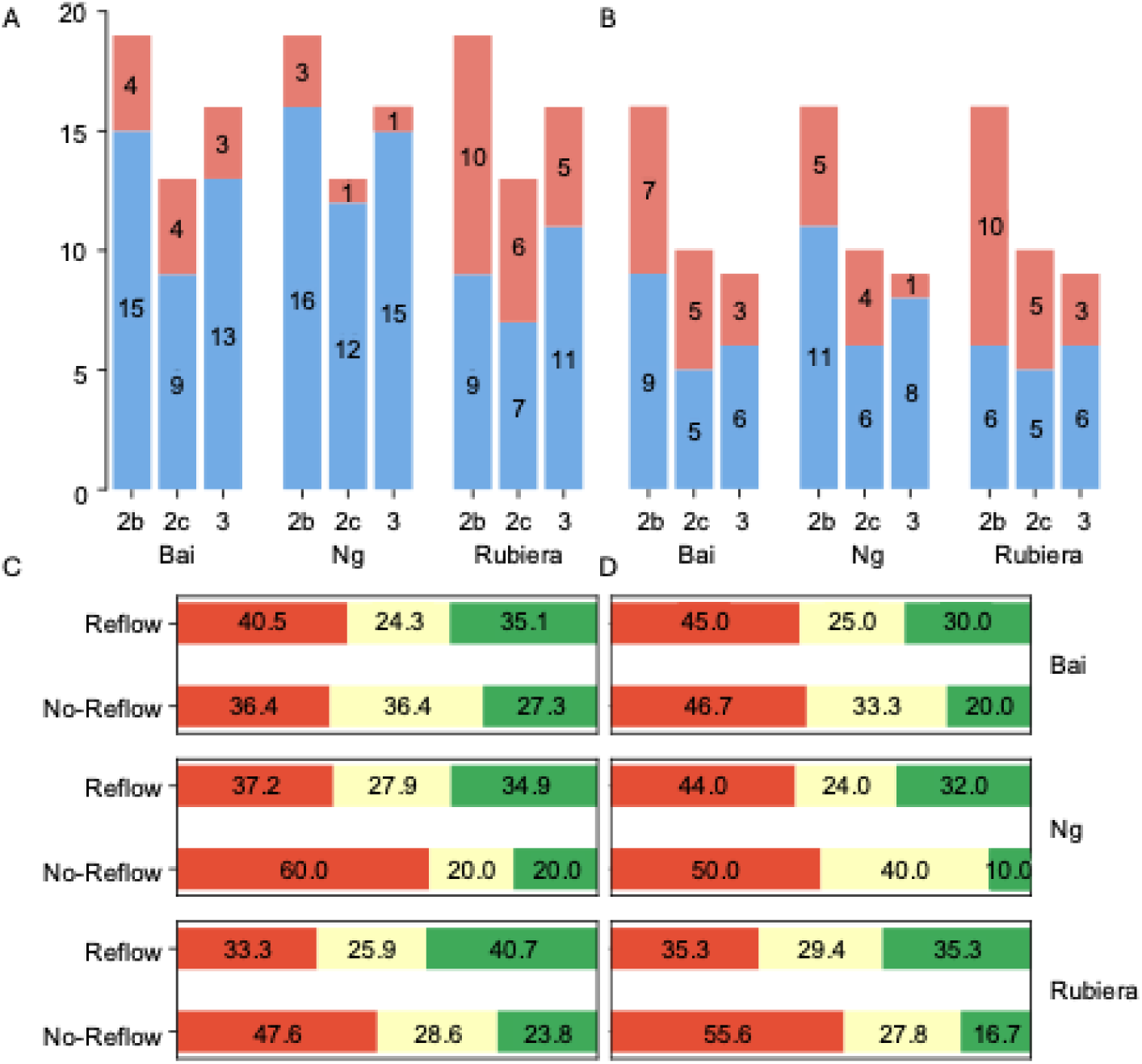
A) No-reflow counts by method for the early (B) and follow-up cohorts (C). mTICI score distribution for the early (C) and late (D) cohorts. Red corresponds to mTICI 2b, yellow 2c and green 3. There is a consistent increase in no-reflow counts for all methods and mTICI scores in the follow-up scan cohort. mTICI 2b is the most represented class for no-reflow across all methods and cohorts.

### Dependence on Time to Follow-Up Imaging

For all methods, no-reflow frequency is larger in the follow-up cohort than that of the early scan cohort. In the intersect cohort, the number of no-reflow cases increases in the follow-up timepoint for all three methods. The method of Bai is the most temporally consistent with a Cohen’s kappa agreement of .296 between the early and follow-up timepoints of the intersect cohort. Ng’s method was most sensitive to the timing of post-EVT imaging, with a Cohen’s kappa of 0.109 between the early and follow-up timepoints. Rubiera’s method shows slightly less temporal consistency than Bai*’*s with a kappa score of 0.203.

## Discussion

Our work reinforces the conclusions of previous studies by demonstrating that the methods in this analysis effectively characterize post-EVT perfusion abnormalities in the cerebral parenchyma. However, because the hemodynamics of microvascular structures cannot be directly observed, as in the case of cardiac no-reflow, we can only reasonably infer micro-circulatory complications when there is persistent parenchymal hypoperfusion after intervention *without alternative explanation*. Therefore, in order to ascertain which methods provide the most objective signal for no-reflow, we must determine which of these methods is least likely to present hypoperfusion caused by confounding processes. From the results of our analysis, we can observe the methods of Rubiera and Bai may be sensitive to factors other than no-reflow. Our results also demonstrate that timing and recanalization threshold also influence no-reflow assessment methods uniformly.

Recanalization quality at mTICI 2b likely influences perfusion parameters sufficiently to preclude the inferral of microcirculatory complications. Across all methods, no-reflow occurrence rates drops by roughly 50% when we removed mTICI 2b cases from consideration. This observed decline may serve to explain the disparity in no-reflow occurrence rates between Mutimer and the originally reported rates, as their analysis only made use of patients with mTICI 2c or better. Notably, the drop in no-reflow occurrence is still present in the early cohort applied to Rubiera ‘s method, which is designed for imaging taken much earlier the 24hr follow-up time of Mutimer ‘s cohort and the cohorts of all other methods.

Rubiera ‘s method is sensitive to baseline hypoperfusion volume and may be unreliable for strokes with medium to large pre-EVT ischemic areas. Rubiera et. al. do not explicitly study no-reflow. However, they find that a hypoperfusion volume (Tmax > 6 s) < *3*.*5*cc predicts both dramatic clinical recovery, defined as NIHSS of ≤ *2* or a ≥ *8* point drop in NIHSS at 24 hours post-intervention, and favorable outcome. Therefore, when adopting this imaging profile as a proxy for no-reflow, one imposes a uniform threshold of 3.5 cc. The change over time of the volume of Tmax > 6 s lesion likely contains signal for perfusion complications or no-reflow.

However, a uniform threshold on the dynamics of hypoperfusion volume may not be appropriate or accurate for identifying no-reflow. In particular, for larger strokes, the core volume of infarcted tissue will be larger and possibly exceed the 3.5 cc threshold, even if all ischemic tissue is adequately reperfused. Our results lend credence to this notion, as all stroke cases with pre-intervention hypoperfusion volumes larger than 130 cc have hypoperfusion volumes larger than 3.5 cc after EVT across both cohorts. Moreover, we observe a statistically significant reduction in mean hypoperfusion volume between reflow and no-reflow cohorts as determined by Rubiera ‘s method as illustrated in Figure 4.

**Figure 4:**
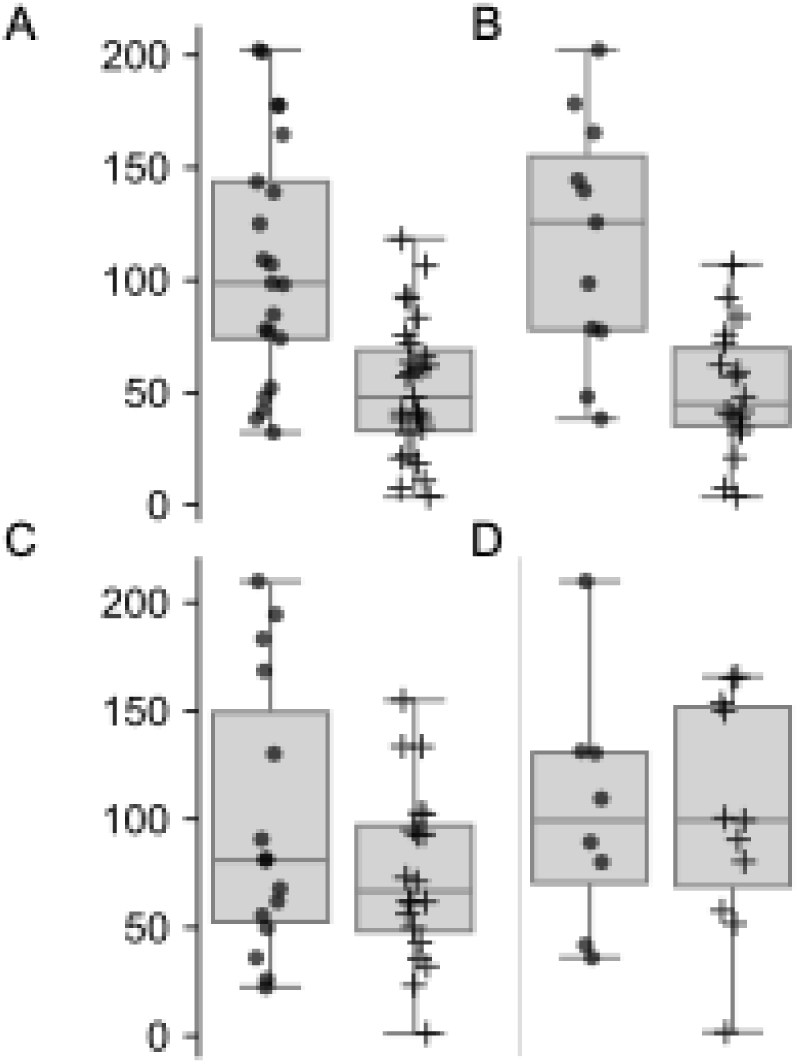
Box and whisker plots of pre-EVT Tmax > 6 s lesion volumes grouped by no-reflow status according to Rubiera ‘s definition for the early (A) and follow-up (B) cohorts. Dots represent reflow cases and pluses represent no-reflow cases. For both cohorts, there is no overlap between the inter-quartile ranges. Welch’s one-sided t-test shows that the mean of the no-reflow Tmax > 6 s lesion volume is smaller than that of reflow cases with rejection of the null hypothesis (p values 0.0009 and 0.004, respectively). (C) and (D) represent Tmax > 6 s lesion volumes grouped by no-reflow status according to Bai ‘s definition for the early and follow-up cohorts, respectively. With a similar statistical testing setup, there is no significant difference in the mean Tmax > 6 s volumes.

Bai’s method may provide a path towards more accurately inferring micro-circulatory status from the temporal dynamics of Tmax > 6 s volume. There are no statistically significant differences in the means of the pre-treatment Tmax > 6 s volumes between reflow and no-reflow subcohorts determined by Bai ‘s method. One potential explanation for the lack of such a relationship is the choice of a relative threshold for TOR, which is informed by final functional independence results from previous studies. There are still some possible confounders, however. For example, a significant delay between baseline imaging and EVT (on the order of 60-120 minutes or more) can allow substantial ischemic core growth [41]. In fast infarct progression cases, this delay could potentially increase ischemic core past the 10% threshold of the Tmax > 6 s lesion, classifying a case as no-reflow even if all ischemic tissue was successfully reperfused with no microcirculatory issues afterwards.

Ng’s method exhibits higher sensitivity to time since EVT than the methods of Rubiera and Bai. Ng ‘s method uses post-treatment DWI to determine the ischemic lesion to determine no-reflow. Moreover, the volume of that lesion does not influence the final no-reflow determination as directly as the methods of Rubiera and Bai. For this method, no-reflow occurrence more than triples for the follow-up cohort, in which post-EVT treatment is taken much closer to the standard 24 hour imaging schedule. Similarly, Bai ‘s method is closest to originally reported frequency at follow-up, which is also closer to Bai ‘s originally reported range of 24 to 72 hrs. As such, it seems that these methods are more sensitive to no-reflow when time since EVT is closer to that of the original literature.

The time dependence of each method poses some questions for further consideration. In particular, because the quantitative perfusion imaging used in this and other analyses can only serves as a proxy for microscopic hemodynamics, we must question whether or not the variance in no-reflow rates with respect to time since EVT accurately characterizes the temporal dynamics of no-reflow. This is especially important to consider for further investigation, as physicians will receive more benefit from imaging profiles that can detect no-reflow when it can be reliably detected within the intervention window. The results of our analysis support the notion that no-reflow occurs after some delay after intervention, at least at a clinically detectable scale.

Our analysis allows us to comment on whether or not physicians can infer micro-circulatory complications from quantitative perfusion imaging. However, none of the imaging profiles provide a measure of the severity of no-reflow. It is possible that no-reflow occurs immediately after EVT and cannot be detected until it reaches a certain severity, which may not occur until several hours after the intervention window. (One potential approach for further strengthening the ability to infer perfusion abnormality instead of just infarct core growth is to differentiate between tissue that is ischemic and tissue that is infarcted on the perfusion parameter maps.)

There are weaknesses to the analysis we have conducted. Namely, we have a relatively small number of patients for our analysis, and a follow-up study with more patients would lend more statistical power to the results. Another weakness is that we do not consider CT patients (patients that underwent CT perfusion imaging rather than MR) or mixture patients (either the pre-EVT is CT perfusion and post-EVT is MR perfusion or vice-versa), limiting the applicability of our results. Another weakness of our paper is that we use the mTICI score that is reported by physicians during the procedure, whereas other studies use ‘core laboratory adjudicated angiographic assessment’, which is more robust to variability attributable to individual operators [24].

## Conclusion

We present an exploratory analysis of the influence on time since EVT on no-reflow detection rates across multiple published assessment methods. Our findings show significant variability in no-reflow detection across all studied methods, with rates ranging from 4.2% to 51.4% depending on method, timing of follow-up imaging, and recanalization quality thresholds. Time since EVT substantially influences no-reflow detection rates across all methods, with follow-up scans (< *48* hours post-EVT) producing rates more consistent with original literature compared to early scans (< *24* hours post-EVT). This temporal dependence suggests either that microcirculatory complications develop over time or that imaging-based proxies for no-reflow become more apparent with delay. Inclusion of mTICI 2b patients may significantly inflate no-reflow estimates, with rates dropping by approximately 50% when restricting analysis to mTICI 2c/3 cases. This finding may explain discrepancies between studies and suggests that recanalization quality confounds no-reflow assessment. Method agreement varies considerably, with Tmax-based approaches (Bai and Rubiera) showing higher concordance than comparisons involving the rCBV/rCBF method of Ng, particularly in the early cohort. The substantial temporal dependence across all methods raises questions about optimal imaging timing for clinical decision-making, as detection may occur outside the therapeutic window for intervention. Opportunities for future work include refining methods to enable detection no-reflow within the intervention window. Further work is also needed to clarify the influence of recanalization quality and other baseline characteristics that confound no-reflow assessment, such as collateral flow.

## Data Availability

Due to HIPAA concerns, data cannot be shared without IRB approval.

## Sources of Funding

This research is supported by NIH National Institute of Neurological Disorders and Stroke under Award R01NS100806.

## Disclosures

None

## Non-standard Abbreviations and Acronyms

AIS: acute ischemic stroke
EVT: endovascular thrombectomy
LVO: large vessel occlusion
TICI: thrombolysis in cerebral infarction
mTICI: modified thrombolysis in cerebral infarction
eTICI: thrombolysis in cerebral infarction
mRS: modified rankin score
DSA: digital subtraction angiography
CT: computed tomography
MR: magnetic resonance
rCBV: relative cerebral blood volume
rCBF: relative cerebral blood flow
MTT: mean transit time
Tmax: time to maximum of the residue function
TOR: tissue optimal reperfusion
HT: hemorrhagic transformation
DWI: diffusion weighted imaging
FLAIR: fluid-attenuated inversion recovery
DSC-MRP: dynamic susceptibility contrast magnetic resonance perfusion imaging
GRE: gradient echo
ASL: arterial spin labeling
ADC: apparent diffusion coefficient
FMRIB: oxford centre for functional magnetic resonance imaging of the brain
FSL: FMRIB software library
FLIRT: FSL linear registration tool
ROI: region of interest

## Notes

### Competing Interest Statement

The authors have declared no competing interest.

### Author Declarations

The study data was collected and de-identified with approval from the UCLA's institutional review board IRB-18-0329. No informed consent was required.

